# Correlates of nucleocapsid antibodies and a combination of spike and nucleocapsid antibodies against protection of SARS-CoV-2 infection during Omicron XBB.1.16 and EG.5 predominant wave

**DOI:** 10.1101/2024.02.21.24303122

**Authors:** Shohei Yamamoto, Yusuke Oshiro, Natsumi Inamura, Takashi Nemoto, Tomofumi Tan, Kumi Horii, Kaori Okudera, Maki Konishi, Tetsuya Mizoue, Haruhito Sugiyama, Nobuyoshi Aoyanagi, Wataru Sugiura, Norio Ohmagari

**Affiliations:** Department of Epidemiology and Prevention, Center for Clinical Sciences, National Center for Global Health and Medicine, Tokyo, Japan; Department of Laboratory Testing, Center Hospital of the National Center for the Global Health and Medicine, Tokyo, Japan; Infection Control Office, Center Hospital of the National Center for the Global Health and Medicine, Tokyo, Japan; Infection Control Office, Kohnodai Hospital of the National Center for the Global Health and Medicine, Chiba, Japan; Center Hospital of the National Center for the Global Health and Medicine, Tokyo, Japan; Kohnodai Hospital of the National Center for the Global Health and Medicine, Chiba, Japan; Center for Clinical Sciences, National Center for Global Health and Medicine, Tokyo, Japan; Disease Control and Prevention Center, National Center for Global Health and Medicine, Tokyo, Japan

## Abstract

**Background:** The role of nucleocapsid (N) antibodies and their combination with spike (S) antibodies against SARS-CoV-2 reinfection remains unclear. We aimed to examine the association between N antibodies, a combination of N and S antibodies, and protection against SARS-CoV-2 reinfection.

**Methods:** We conducted a prospective cohort study among staff at a national medical research center in Tokyo and followed them for the incidence of SARS-CoV-2 infection between June and September 2023 (Omicron XBB.1.16/EG.5 predominant wave). At baseline, participants donated blood samples to measure N-and S-specific antibodies in assays from three companies (Roche, Abbott, and Sysmex). Cox regression was used to estimate the hazard ratio (HR) and protection (1-HR*100) against subsequent SARS-CoV-2 infection across these antibody levels.

**Findings:** Of the 2549 staff included in the analysis, 237 SARS-CoV-2 infections were identified during follow-up. Among participants with previous infection, higher pre-reinfection N antibodies were associated with a lower risk of reinfection even after adjusting S antibody levels (P for trend<0.01). Estimation of the protection matrix for N and S antibodies yielded that high levels in both N and S antibodies conferred robust protection (>90%) against subsequent infection. In addition, a pattern of low pre-reinfection N antibodies but high vaccine-enhanced S antibodies showed high protection (>80%).

**Interpretation:** Pre-reinfection N antibody levels correlated with protection against reinfection, independent of S antibodies. If the N antibodies were low, vaccine-boosted S antibodies could enhance the reinfection protection.

**Funding:** National Center for Global Health and Medicine and Japan Health Research Promotion Bureau Research Fund.

**Research in context:** 

**Evidence before this study:** We searched published and preprinted literature with the following keywords: “COVID-19,” “SARS-CoV-2,” “nucleocapsid,” “spike,” “antibody,” “protection,” and “reinfection.” We found few prospective or case-control studies examining the association between pre-reinfection anti-SARS-CoV-2 nucleocapsid (N) antibody levels and risk of SARS-CoV-2 reinfection; in particular, no studies were conducted for adults among Omicron-dominant phases. We also found no studies that examined the role of a combination of anti-spike (S) and anti-N antibodies in protection against SARS-CoV-2 infection.

**Added value of this study:** This study first revealed that pre-reinfection anti-N antibody levels correlated with protection against reinfection during the Omicron XBB.1.16 and EG.5 predominant waves even after adjusting S antibody levels. Further, we first estimated the protection matrix by combining anti-N and S antibody levels and showed that both high levels in N and S conferred robust protection (>90%). Vaccine-induced higher S antibody levels were associated with higher protection among previously infected individuals with low levels of N antibodies.

**Implications of all the available evidence:** The prolonged COVID-19 pandemic has resulted in diverse immune characteristics across individuals due to varying timing of infection and doses and timing of vaccination, making it challenging to decide the timing of additional vaccination. Our results suggest the utility of assessing both N and S antibody levels for considering the timing of additional vaccination for those with a history of COVID-19. If the N antibody level was low due to waning over time, additional vaccination enhances S antibodies and might improve the protection against reinfection.

## Introduction

Four years into the COVID-19 pandemic, over 774 million COVID-19 cases have been reported worldwide as of January 2024.^1^ Currently, reinfection with the new variant has become an important public health concern, underscoring the need to identify predictors for prevention. Previous SARS-CoV-2 infection is associated with a lower risk of reinfection.^2^ Among antigen-specific immune responses elicited during SARS-CoV-2, spike (S) antibodies are known to have a role in preventing virus entry, and epidemiological studies confirmed that higher S antibodies correlated with higher protection against reinfection.^3,4^ In contrast, the protective role of other antigen-specific immune responses remains unclear.

Nucleocapsid (N)-specific antibodies have been used as a marker of previous infection. Interestingly, in vivo studies showed that N antibodies elicited antibody-dependent cellular cytotoxicity (ADCC) effects^5^ and correlated with protection against the SARS-CoV-2 challenge.^5–7^ Data from human epidemiological studies are scarce and inconsistent. In a study among children with a history of COVID-19, higher N titers were associated with a significantly lower risk of reinfection during the Omicron BA.4/5 phase (2022).^8^ In a study of male adults, higher N titers were marginally associated with a lower risk of reinfection during the Delta predominant wave (2021).^9^ In contrast, two other studies among adults with a small sample size (less than 120 individuals) reported no association between initial infection-acquired N antibody levels and a risk of reinfection during the early pandemic phase (2020).^3,10^

Besides the paucity of epidemiological data linking N antibody levels to reinfection risk, some issues remain to be addressed. First, no studies controlled for S antibody levels when analyzing the association between N titer and reinfection risk. Because both S and N antibodies are induced by infection and thus highly correlated, the lack of adjustment of S antibodies is critical when assessing the independent role of N antibodies. Second, no evidence is available in adults during the surge of the Omicron variants, wherein many infections and reinfections have occurred. Third, no studies investigated the combination of S and N antibody levels in relation to the risk of reinfection. In vivo studies showed that mice with both S and N antibodies had better protection than those with either S antibodies only,^11–13^ suggesting that higher levels in both S and N antibodies may confer robust protection.

Here, we studied the association of pre-reinfection N antibody level and a combination of S and N antibodies with the risk of SARS-CoV-2 reinfection during the Omicron XBB.1.16/EG.5 predominant wave among the staff of a national medical and research center in Tokyo.

## Methods

### Study Setting

In the National Center for Global Health and Medicine (NCGM) in Japan, a repeat serological study was launched in July 2020 to monitor the spread of SARS-CoV-2 infection among staff during the COVID-19 epidemic. The details of this study have been reported elsewhere.^14,15^ In summary, we have completed eight surveys as of June 2023. We measured anti-SARS-CoV-2 N-(all surveys) and S-protein antibodies (from the second survey onward) for all the participants using Roche and Abbott assays and stored serum samples at −80°C. We also measured N and S antibodies with Sysmex assays in four surveys (once a year). In addition, we collected information on COVID-19–related factors, including vaccination, occupational infection risk, and infection prevention practices via a questionnaire. The self-reported vaccination status was validated against the vaccine records owned by the NCGM Labor Office. Written informed consent was obtained from all the participants. This study was approved by the NCGM Ethics Committee (approval number: NCGM-G-003598).

### Analytic cohort

In the present prospective study, we set the baseline cohort as all participants who attended the eighth survey conducted in June 2023, where we invited all NCGM staff (n=3206), and 2569 (80%) completed a questionnaire and donated blood samples. Of those, we excluded 20 participants who lacked information on covariates: body composition (n=12), alcohol drinking status (n=3), living arrangement status (n=6), adherence to infection prevention practice (n=5), and infection risk behaviors (n=3); thus, 2549 participants were analyzed.

### Ascertainment of subsequent SARS-CoV-2 infection

We followed the participants for COVID-19 incidence using the COVID-19 patient records documented by the NCGM Hospital Infection Prevention and Control Unit, which provided information on the date of diagnosis, diagnostic procedures, symptoms, and hospitalizations. As per the NCGM rule, staff should undergo PCR or antigen test for COVID-19 when they have COVID-19-compatible symptoms, and if it tests positive, they must report the results to the NCGM Hospital Infection Prevention and Control Unit. Most registered cases were laboratory-confirmed (PCR or antigen test), but there were a few exceptions, including those diagnosed by a physician based on non-laboratory information (i.e., symptoms compatible with COVID-19 after close contact with a patient with COVID-19).

In this analysis, we defined reinfection as being diagnosed more than 90 days after a previous diagnosis, according to the Centers for Disease Control and Prevention (CDC).^16^ However, no cases were re-diagnosed within 90 days after the previous infection; the minimum interval was 227 days. We also considered the following pattern as reinfection: individuals who were first diagnosed with COVID-19 during follow-up but had a history of seropositive N antibodies at baseline.

### Antibody testing

We assessed anti-SARS-CoV-2 N and S protein antibodies in all the participants. N antibody was measured using three commercially available automated immunoassays: (1) Elecsys^®^ Anti-SARS-CoV-2, Roche Diagnostics; (2) ARCHITECT SARS-CoV-2 IgG, Abbott Laboratories; and (3) HISCL SARS-CoV-2 N-IgG, Sysmex Co. The antibodies measured with Roche (cut-off index: COI) and Abbott (signal to cut-off: S/CO) assays are qualitative, while those with Sysmex (Sysmex unit: SU/mL) are quantitative. Abbott and Sysmex assays measure IgG N, whereas Roche assay measures total N including IgG. We also quantitatively measured the antibodies against the receptor-binding domain (RBD) of the SARS-CoV-2 S protein using the Elecsys^®^ Anti-SARS-CoV-2 S RUO, Roche Diagnostics (i.e., anti-RBD total, U/mL) and the AdviseDx SARS-CoV-2 IgG II assay, Abbott Laboratories (i.e., anti-RBD IgG, AU/mL) and that against the SARS-CoV-2 IgG S protein using HISCL SARS-CoV-2 S-IgG, Sysmex Co. (i.e., anti-S IgG, SU/mL).

### Previous SARS-CoV-2 infection status at baseline

Previous infection was defined as a self-reported history of COVID-19 (confirmed against in-house COVID-19 registry) at baseline or anti-N seropositive with any of the three assays (Roche ≥1.0 COI, Abbott ≥ 1.40 S/C, or Sysmex ≥10 SU/mL) at any of the first (July 2020) through eighth (June 2023: baseline) surveys. Participants were dichotomized based on infection status at baseline: infection-naïve or previously infected. The latter was further divided into quartile groups according to the N antibody level on each of the three assays.

### Statistical analysis

We calculated the person-time from the date of the baseline blood sampling (June 13–23, 2023) to the date of subsequent SARS-CoV-2 infection, receiving an additional COVID-19 vaccine, or censoring (September 6, 2023), whichever occurred first. We fitted a Cox proportional hazard regression analysis to examine the association between N antibody status (i.e., infection-naïve group and N index quartile groups of previously infected) and the risk of subsequent SARS-CoV-2 infection during the Omicron BBX.1.16/EG.5 predominant wave for each of N antibodies measured with three companies. Models were adjusted in the following manner. Model 1 was adjusted for age and sex. Model 2 was additionally adjusted for job, occupational SARS-CoV-2 exposure risk, body mass index, comorbid diseases, immunosuppression, use of tobacco products, frequency of alcohol drinking, number of households, number of live-in school-aged children, infection prevention practice score, frequency of spending ≥30 min in the 3Cs (crowded places, close-contact settings, and confined and enclosed spaces) without mask, and frequency of having dinner in a group of ≥5 people for >1 h. Model 3 was further adjusted for anti-S/RBD titer measured with the assay of the same company.

Among the previously infected individuals, we examined the association between the N antibody index and the risk of SARS-CoV-2 reinfection using restricted cubic splines with three knots at the 10th, 50th, and 90th centiles of the N antibody distribution, which based on the Cox proportional regression analysis with adjustment for the covariates of Model 2.

To examine the association between a combination of S and N antibody levels and protection against SARS-CoV-2 infection, we repeated the Cox regression model with adjustment for covariates of Model 2. We used combined variables for each category of N antibody status (i.e., infection-naïve group and N index quartile groups of previously infected) and S antibody status (i.e., quartile groups) and set the reference group as infection-naïve and the lowest quartile of S antibodies. Further, we fitted the Cox model while accounting for continuous S and log-N antibodies as an interaction term, and the results of this analysis were conveyed visually in contour plots. In this contour plots model, the minimum values of S and N antibodies were selected as reference values. The estimated hazard ratio (HR) was used to calculate protection (%), according to the formula: (1 – HR) × 100.

As a sensitivity analysis, we repeated the above analyses by restricting the outcome to symptomatic infection. As another sensitivity analysis, we run the analyses after excluding non-regular staff (i.e, contractors, temporary staff, café staff, shop staff, and part-time registered medical doctors) since the infection for non-regular staff might not be completely reported to the NCGM registry. Statistical analyses were performed using Stata 18.0 (StataCorp LLC). All *P* values were two-sided, and P < 0.05 was considered statistically significant.

## Results

### Baseline characteristics

Of 2549 participants, 70.9% were female, and the median age was 38 years (**Table 1**). The most frequent jobs were nurses (35.9%), followed by doctors (16.0%), allied healthcare workers (15.4%), administrative staff (15.2%), and researchers (12.2%). More than half (56%) were previously infected with SARS-CoV-2; of those, 29% had no history of COVID-19 diagnosis but with a history of seropositive on N antibodies at baseline, and 8.7% had a history of COVID-19 diagnosis but no history of seronegative on N antibodies at baseline. Baseline characteristics stratified by previous infection status and Roche N index were summarized in **Table 1**. Previously infected individuals tended to be younger, at higher risk of occupational exposure to SARS-CoV-2, have fewer comorbidities, drink alcohol more frequently, engage in high-risk behaviors, live with more school-aged children, receive fewer doses of the COVID-19 vaccine, and have fewer receiver of the Omicron bivalent (BA.1 or BA.4/5) vaccines compared with infection-naïve individuals. Among those with previous infection, a higher Roche N index was correlated with a shorter interval from the last COVID-19 diagnosis to the baseline and a higher number of experienced COVID-19 diagnoses.

**Table 1.** Baseline characteristics according to anti-SARS-CoV-2 nucleocapsid index.

| Characteristics | Total | Infection-naïve | Quartile of Roche N index among previously infected individuals |  |  |  |
| --- | --- | --- | --- | --- | --- | --- |
|  |  |  | Q1 (lowest) | Q2 | Q3 | Q4 (highest) |
| Participants | N=2,549 | N=1,119 | N=358 | N=357 | N=358 | N=357 |
| Female, % | 70.9 | 73.0 | 70.4 | 68.3 | 69.3 | 68.6 |
| Age, y | 38 (27–49) | 41 (29–53) | 38 (28–47) | 33 (26–45) | 34 (27–46) | 35 (27–48) |
| Job, % |  |  |  |  |  |  |
| Doctor | 16.0 | 11.6 | 22.1 | 18.2 | 17.0 | 20.4 |
| Nurse | 35.9 | 33.1 | 40.2 | 40.6 | 36.6 | 34.7 |
| Allied healthcare worker | 15.4 | 17.9 | 10.3 | 14.6 | 15.9 | 13.2 |
| Researcher | 12.2 | 13.5 | 10.3 | 9.0 | 12.8 | 12.3 |
| Administrative staff | 15.2 | 18.4 | 12.0 | 11.5 | 12.3 | 14.8 |
| Others | 5.4 | 5.5 | 5.0 | 6.2 | 5.3 | 4.5 |
| Occupational SARS-CoV-2 exposure risk <sup>a</sup>, % |  |  |  |  |  |  |
| Low | 61.2 | 65.1 | 60.1 | 56.0 | 60.3 | 56.3 |
| Moderate | 20.7 | 19.7 | 18.4 | 23.2 | 20.9 | 23.0 |
| High | 18.1 | 15.1 | 21.5 | 20.7 | 18.7 | 20.7 |
| Body mass index, kg/m<sup>2</sup> | 21.2 (19.5–23.4) | 21.1 (19.4–23.6) | 21.2 (19.7–23.3) | 21.1 (19.4–23.0) | 21.2 (19.5–22.9) | 21.3 (19.7–23.5) |
| Comorbid diseases <sup>b</sup>, % | 8.3 | 10.5 | 8.7 | 4.8 | 6.4 | 6.7 |
| Immunosuppression <sup>c</sup>, % | 1.0 | 1.0 | 0.8 | 0.6 | 1.7 | 0.8 |
| Use of tobacco products <sup>d</sup>, % | 7.6 | 8.3 | 5.6 | 8.4 | 7.0 | 7.3 |
| Frequency of alcohol drinking, % |  |  |  |  |  |  |
| None | 32.9 | 37.2 | 30.7 | 28.6 | 31.6 | 27.5 |
| Occasional | 27.1 | 25.8 | 26.3 | 28.3 | 27.7 | 30.0 |
| Weekly/Daily | 40.0 | 37.0 | 43.0 | 43.1 | 40.8 | 42.6 |
| No. of households | 2 (1–3) | 2 (1–3) | 2 (1–4) | 2 (1–4) | 2 (1–4) | 2 (1–4) |
| No. of school-aged children cohabiting <sup>e</sup>, % |  |  |  |  |  |  |
| 0 | 71.0 | 77.3 | 59.5 | 70.6 | 64.5 | 69.5 |
| 1 | 13.0 | 12.4 | 14.0 | 12.0 | 14.5 | 13.4 |
| ≥2 | 16.0 | 10.3 | 26.5 | 17.4 | 20.9 | 17.1 |
| Infection prevention practice score <sup>f</sup> | 7 (6–9) | 7 (6–9) | 7 (5–8) | 7 (5–8) | 7 (6–8) | 7 (6–8) |
| Spending ≥30min in the 3Cs without mask, % |  |  |  |  |  |  |
| None | 62.0 | 67.6 | 54.2 | 58.8 | 60.3 | 57.1 |
| 1–5 times | 29.1 | 27.5 | 33.5 | 29.4 | 29.1 | 29.4 |
| ≥6 times | 8.9 | 4.8 | 12.3 | 11.8 | 10.6 | 13.4 |
| Having dinner in a group of ≥5 people for >1h, % |  |  |  |  |  |  |
| None | 58.6 | 64.8 | 50.8 | 56.3 | 53.9 | 53.8 |
| 1–5 times | 37.2 | 32.8 | 42.7 | 39.2 | 42.2 | 38.4 |
| ≥6 times | 4.2 | 2.4 | 6.4 | 4.5 | 3.9 | 7.8 |
| No. of previous COVID-19 diagnosis, % |  |  |  |  |  |  |
| Never | 60.1 | 100.0 | 33.2 | 30.5 | 22.6 | 29.4 |
| 1 | 38.4 | 0 | 65.9 | 69.5 | 75.4 | 63.0 |
| 2 | 1.4 | 0 | 0.8 | 0.0 | 1.7 | 7.3 |
| 3 | 0.1 | 0 | 0.0 | 0.0 | 0.3 | 0.3 |
| Interval from last diagnosis to baseline, d | 279 (183–342) | NA | 332 (280–463) | 309 (206–354) | 239 (178–317) | 190 (112–294) |
| Vaccination status, % |  |  |  |  |  |  |
| ≤2-dose | 4.3 | 4.0 | 4.5 | 4.2 | 5.3 | 4.2 |
| 3-dose | 21.6 | 14.9 | 24.6 | 26.3 | 30.2 | 26.3 |
| 4-dose | 41.8 | 39.1 | 43.9 | 46.2 | 41.1 | 44.5 |
| ≥5-dose | 32.2 | 41.9 | 27.1 | 23.2 | 23.5 | 24.9 |
| Receiver of Omicron bivalent vaccine, % | 47.2 | 55.4 | 46.4 | 42.3 | 38.0 | 36.1 |
| Interval from last vaccination to baseline, d | 255 (181–306) | 214 (178–299) | 275 (182–457) | 286 (182–455) | 294 (186–461) | 293 (186–452) |

Data are presented as median (interquartile range) for continuous variables and percentage for categorical variables.
<sup>a</sup> Occupational SARS-CoV-2 exposure risk was categorized as low (those not engaged in COVID-19–related work), moderate (those engaged in COVID-19–related work without heavy exposure to SARS-CoV-2), or high (those heavily exposed to SARS-CoV-2).
<sup>b</sup> Comorbid diseases were defined as cancer, cardiovascular disease, diabetes, hypertension, chronic kidney disease, or lung disease.
Tobacco products include conventional cigarettes and heated tobacco products.
<sup>c</sup> Immunosuppression was defined as having an immunosuppressive disease or using steroids [except topical or inhaled], immunosuppressants, or anticancer drugs
<sup>d</sup> Tobacco products include conventional cigarettes and heated tobacco products.
<sup>e</sup> School-age children include those in nurseries, kindergartens, elementary to high school, university, and with disabilities
<sup>f</sup> Infection prevention practice score was calculated on the basis of the total score of adherences to avoiding the 3Cs, hand washing, wearing a mask, social distancing, and not touching the face, nose, or mouth, assigning 2 points to “always,” 1 point to “often,” and 0 points to others (“seldom” and “not at all”).
*Abbreviations:* 3Cs, crowded places, close-contact settings, and confined and enclosed spaces; COVID-19, coronavirus disease 2019; Q: quartile, SARS-CoV-2, severe acute respiratory syndrome coronavirus 2.

### Incidence of subsequent SARS-CoV-2 infection

During the follow-up, we identified 237 SARS-CoV-2 infections, with an incident rate of 13.5 per 10000 person-day. Of those, 192 were diagnosed for the first time, 42 for the second time, and 3 for the third time. Among those diagnosed for the first time, 17 had a history of seropositive on N antibodies at baseline, indicating they have been infected at least two times.

### Previous infection status, pre-reinfection N antibodies, and subsequent SARS-CoV-2 infection

The infection-naïve group had a higher risk of SARS-CoV-2 infection compared with the previously infected group with the lowest quartile of the Roche N index: the HR (95% CI) of 1.93 (1.32–2.83) in Model 2 (**Table 2**). Within the previously infected groups, a higher pre-reinfection Roche N index was associated with a lower risk of reinfection: the HR (95% CI) from lowest to highest quartile groups were 1.00 (reference), 0.29 (0.14–0.57), 0.31 (0.16–0.60), and 0.14 (0.05–0.35), respectively in Model 2 (P for trend<0.01). The association was still significant after adjusting the anti-RBD titer in Model 3. The cubic spline analysis yielded a similar dose-response curve showing a steady decrease in HRs with a higher pre-reinfection Roche N index (P for linearity<0.01) (**Figure 1**). These results were similar in Abbott and Sysmex assays (**Table 2** & **Figure 1**).

**Figure 1.**
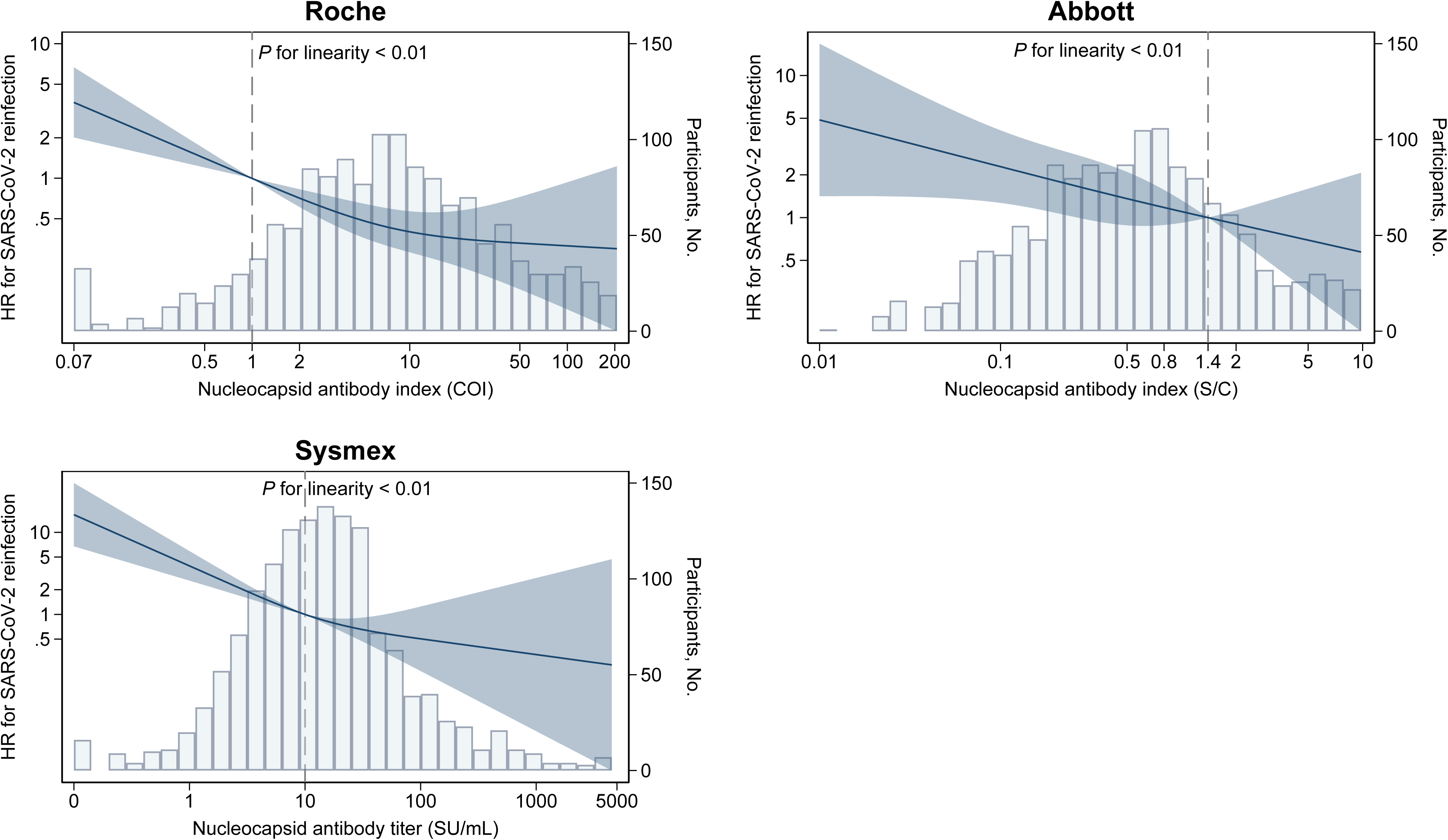
Association between anti-SARS-CoV-2 nucleocapsid antibody level and risk of reinfection among previously infected individuals Solid lines indicate the hazard ratio for SARS-CoV-2 reinfection, and the shaded area represents 95% confidence intervals. The bars indicate histograms of log-transformed nucleocapsid antibody levels. Reference points are seropositive thresholds for each assay (1.0 COI for Roche, 1.4 S/C for Abbott, and 10.0 SU/mL for Sysmex). All models were adjusted for covariates of Model 2 in Table 2. *Abbreviations*: COI, cut-off index; IgG, immunoglobulin G; N, nucleocapsid; Q, quartile; S/CO, signal to cut-off; SU, Sysmex unit.

**Table 2.**
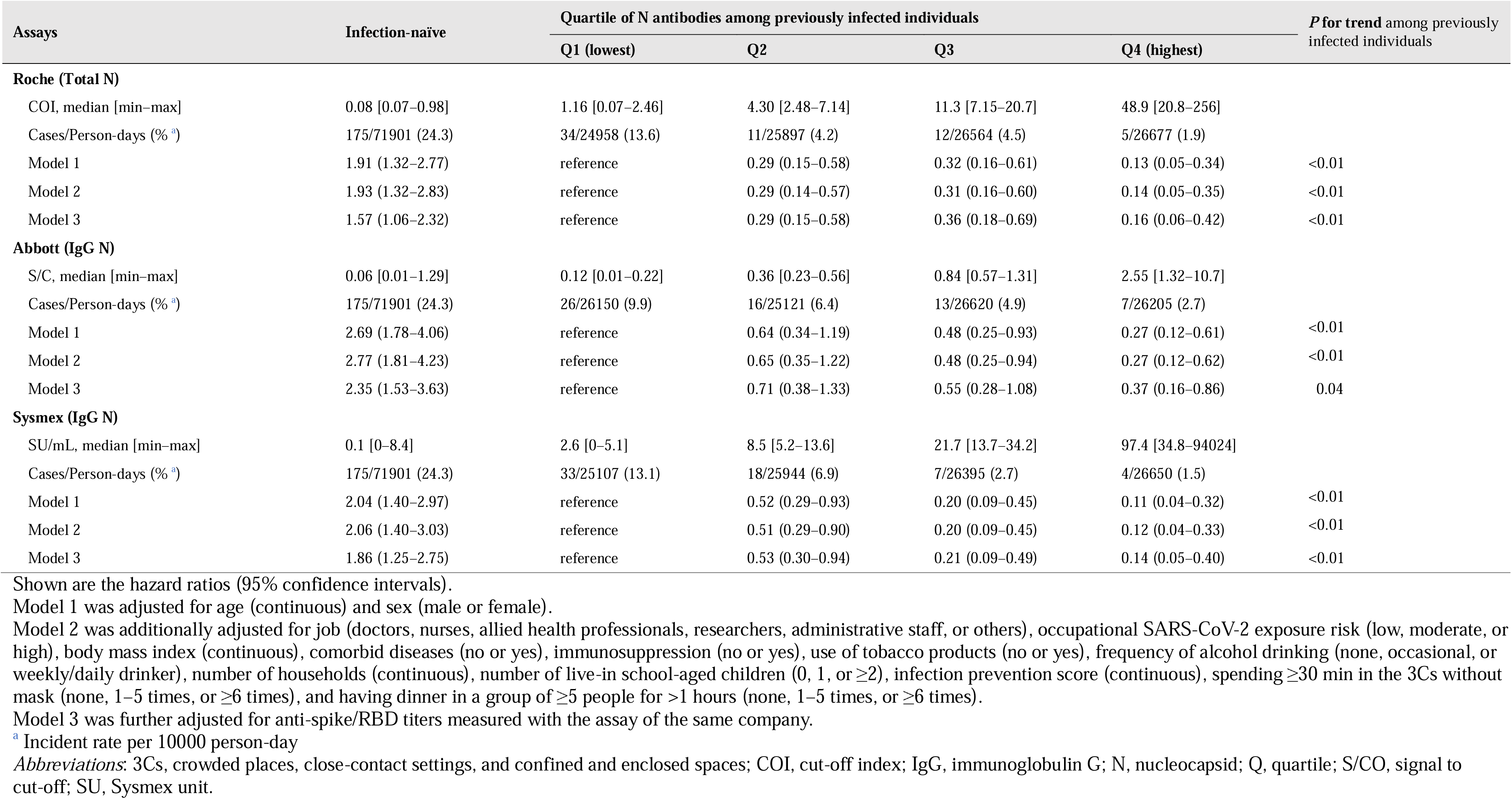
Hazard ratios (95% confidence intervals) for subsequent SARS-CoV-2 infection across the baseline anti-nucleocapsid antibody index.

### Correlations between N and S antibodies

Low correlations were observed between N and S/RBD antibody levels among previously infected individuals: Spearman’s ρ (95% CI) between N and S/RBD antibodies measured with Roche, Abbott, and Sysmex assays were 0.23 (0.18–0.28), 0.32 (0.28–0.37), and 0.34 (0.29–0.38), respectively (**Supplemental Figure 1**). Irrespective of the N antibody levels, participants with greater vaccine doses or shorter intervals from the last vaccination had higher levels of anti-RBD antibodies (**Supplemental Table 1**).

### A joint association of N and S antibodies with protection against subsequent SARS-CoV-2 infection

Figure 2 shows the infection protection matrices for a combination of N and S/RBD antibody levels. The previously infected groups with the highest quartile of the Roche N index had extremely high protection irrespective of RBD antibody levels ranging from 93% to 100%. In the previously infected groups with the lowest quartile of the Roche N index, higher RBD antibody levels were associated with greater protection: the protection (95% CI) from lowest to highest RBD quartile were -35% (-150 to 27), 53% (11 to 75), 71% (39 to 86), and 87% (58 to 96), respectively (P for trend<0.01). In the infection-naïve groups, higher anti-RBD titers correlated only slightly and not significantly, with protection: the protection (95% CI) from the lowest to highest RBD quartile were 0 (reference), 34% (6 to 54), 37% (-1 to 60), and 37% (-17 to 65), respectively (P for trend=0.15). The contour plot of the continuous S and N antibodies showed that high levels in either S or N antibodies conferred relative protection, and high levels in both S and N conferred further robust protection (Figure 2). These results were similar in Abbott and Sysmex assays.

**Figure 2.**
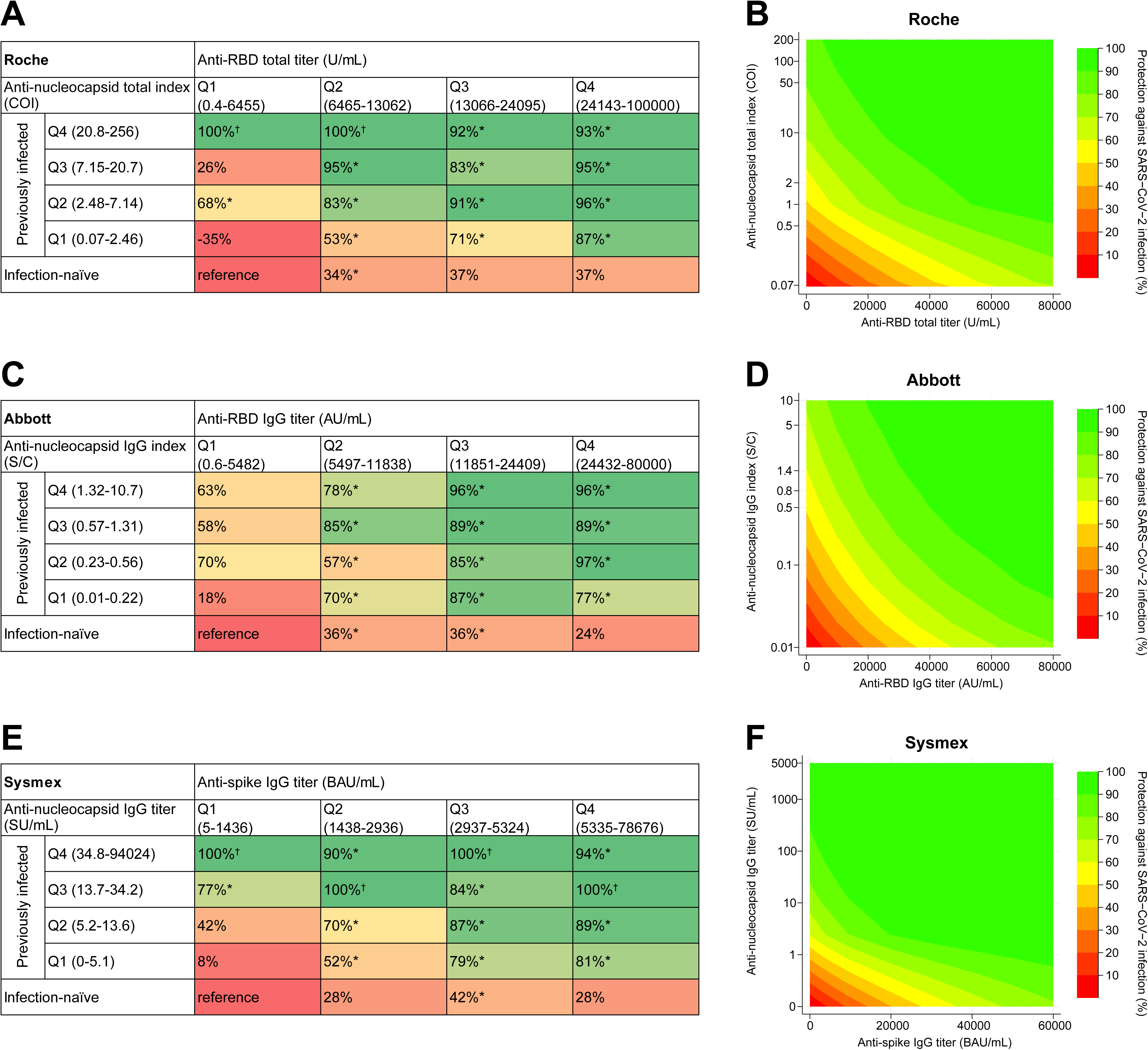
Infection protection matrices for the baseline anti-nucleocapsid and anti-spike/RBD antibody levels Shows are matrix tables of the protection against infection by categorical anti-nucleocapsid and anti-spike/RBD antibodies, relative to the reference group of infection-naïve and lowest quartile of anti-spike/RBD antibodies, with Roche (A), Abbott (C), and Sysmex (E) assays. Also shown are contour plots of the protection by continuous anti-nucleocapsid and anti-spike/RBD antibodies, relative to reference values of the lowest value of anti-nucleocapsid and anti-spike/RBD antibodies, with Roche (B), Abbott (D), and Sysmex (F) assays. Protection was calculated as (1 – hazard ratio) × 100. The hazard ratio was estimated using a Cox proportional hazards regression model, adjusting covariates of Model 2 in Table 2. *: P<0.05 ^†^: No incidence of subsequent SARS-CoV-2 infection in the group *Abbreviations*: AU, arbitrary units; COI, cut-off index; IgG, immunoglobulin G; N, nucleocapsid; Q, quartile; RBD, receptor-binding domain; S/CO, signal to cut-off; SU, Sysmex unit.

## Discussion

During the predominant wave of the Omicron BBX.1.16/EG.5 in Japan, higher levels of pre-reinfection N antibodies were correlated with higher protection against reinfection irrespective of S antibody levels among the previously infected individuals. Those with high levels of both N and S antibodies had robust protection against reinfection. Among those with low levels of infection-acquired N antibodies, higher levels of vaccine-induced S antibodies conferred greater protection against reinfection.

The present finding showing higher protection against reinfection associated with higher pre-reinfection N antibodies among the previously infected individuals agrees with those of two previous studies conducted in Delta and Omicron BA.4/BA.5 predominant phases,^8,9^ but not with the null association reported from other two small studies during earlier pandemic phase.^3,10^ Our study has some important strengths over previous studies, including a larger sample size, well-defined cohort, and identification of COVID-19, details of which will be described later. Additionally, we adjusted for baseline S antibodies, enabling us to assess the independent role of N antibody levels. With these features, our study provides robust evidence of the association between N antibody levels and subsequent risk of reinfection with highly immune evasive Omicron BBX.1.16/EG.5 variants.

The present finding is supported by animal experiments suggesting an independent role of N antibodies against infection. In mouse models, anti-SARS-CoV-2 N antibodies have been shown to improve protection against the SARS-CoV-2 challenge by eliciting natural killer-mediated ADCC against infected cells,^5,17^ which is another critical antibody response for protection besides neutralization.^18^ In mice immunized with an N-specific vaccine, N antibody titer correlated with protection against the SARS-CoV-2 challenge.^6,19^

We first assessed the combined role of N and S antibodies in relation to the risk of SARS-CoV-2 reinfection, showing an extremely high protection rate (>90%) among those high in both N and S antibodies. Animal experiments demonstrated that vaccines encoding both N and S proteins induced higher levels of these antibodies and conferred better protection against SARS-CoV-2 infection than vaccines encoding N or S protein alone,^19,20^ suggesting that enhancing N-specific immunity coupled with S-specific immunity could enhance the protection. In humans, hybrid immunity (i.e., vaccination and infection) shows greater reduction in the risk of SARS-CoV-2 infection than vaccine-induced immunity alone,^2^ which could be partially explained by the joint effect of N and S antibodies.

In previously infected individuals, low N antibody titers represent long time intervals since the infection,^13,21^ asymptomatic or mild symptoms at infection,^22^ or vaccination before the infection.^21,23^ In the present analysis, we found that higher vaccine doses or shorter intervals from the last vaccination were associated with higher S antibodies and conferred greater protection against reinfection among previously infected individuals with low N antibodies. This result emphasizes the importance of additional vaccines for those who were previously infected but have low N antibodies.

During the follow-up period of the present study, the majority (74%) of SARS-CoV-2 infections occurred in infection-naïve participants. Among them, the protection rate showed only 37% even in the highest quartile of Roche RBD titers, indicating a low protective ability of anti-RBD antibodies. The present finding would be reasonable given that anti-RBD antibodies of our infection-naïve participants had been induced by the original monovalent or Omicron bivalent (original + Omicron BA.1 or BA.4/5) vaccines, which has limited capacity to produce neutralizing antibodies against Omicron XXB.1.16 and EG.5.1.^24^ The Omicron XBB.1.5 monovalent vaccination, which started in the autumn of 2023 in several countries, including Japan, is expected to largely reduce the risk of SARS-CoV-2 infection through boosting neutralization capacity against Omicron XXB variants more than the historical vaccines.^25–27^

Strengths of this study include a relatively large size of the cohort, use of antibody titers close to the timing of infection (i.e., short follow-up), measures of N and S antibodies using three types of commercially available antibody assays, comprehensive adjustment for infection risk factors, and rigorous definition of previous and new infection using information on a history of COVID-19 and at most eight-times serological test results since 2020. However, limitations also should be acknowledged. We did not conduct active surveillance of SARS-CoV-2 infection during follow-up, possibly underestimating the number of total infections (e.g., asymptomatic cases).^28^ In the sensitivity analysis restricting only symptomatic cases, we confirmed that the association between N antibody levels and reinfection risk was virtually unchanged (**Supplemental Table 2**). As another limitation, ascertaining SARS-CoV-2 infection by the in-house registry might not be completely recorded for non-regular staff. Nonetheless, we found a similar association between N antibody levels and reinfection risk after excluding non-regular staff (**Supplemental Table 3**).

In conclusion, pre-reinfection N-specific antibody levels correlated with protection against reinfection after controlling for S-specific antibody levels among the healthcare workers during the Omicron XBB.1.16 and EG.5 subvariants dominant wave in Japan. Higher levels of both N and S antibodies conferred robust protection. In previously infected individuals with low levels of N antibodies, vaccine-induced higher S antibody titer enhanced protection against reinfection. The level of N antibodies could have an independent role in infection protection and be a marker for deciding the timing of additional vaccination. Further research is warranted on whether vaccines encoding both S and N proteins confer better protection against infection in humans.

## Data Availability

All data produced in the present study are available upon reasonable request to the authors.

## Contributors

Conceptualization: SY, TM, HS, WS, and NO. Methodology: SY and TM. Software: SY and MK. Validation: MK and TM. Formal analysis: SY. Investigation: SY, YO, NI, TN, TT, KH, KO, MK, and TM. Resources: YO, TM, HS, NA, WS, and NO. Data Curation: SY, MK, and TM. Writing - Original Draft: SY and TM. Visualization: SY. Supervision: TM and NO. Project administration: SY and TM. Funding acquisition: TM. All authors contributed to manuscript revisions and editing. All authors confirm they had full access to data in the study and accept responsibility for submitting it for publishing.

## Data Sharing

The data is not publicly available due to ethical restrictions and participant confidentiality concerns, but de-identified data can be available by contacting the Department of Epidemiology and Prevention, Center for Clinical Sciences, National Center for Global Health and Medicine, Japan (website: http://epid.ncgm.go.jp/, tel: +81 3 3202 7181 [ext 2859]) for the researchers who meet the criteria for access to confidential data.

## Acknowledgment

We thank Mika Shichishima for her contribution to data collection and the staff of the Laboratory Testing Department for their contribution to measuring antibody testing.

## Ethical approval

Written informed consent was obtained from all participants, and the study procedure was approved by the NCGM Ethics Committee (approval number: NCGM-G-003598).

