## Supplemental Materials for "Correlates of nucleocapsid antibodies and a combination of spike and nucleocapsid antibodies against protection of SARS-CoV-2 infection during Omicron XBB.1.16 and EG.5 predominant wave"

**Roche**

Spearman's ρ = 0.23 (95%CI: 0.18-0.28)

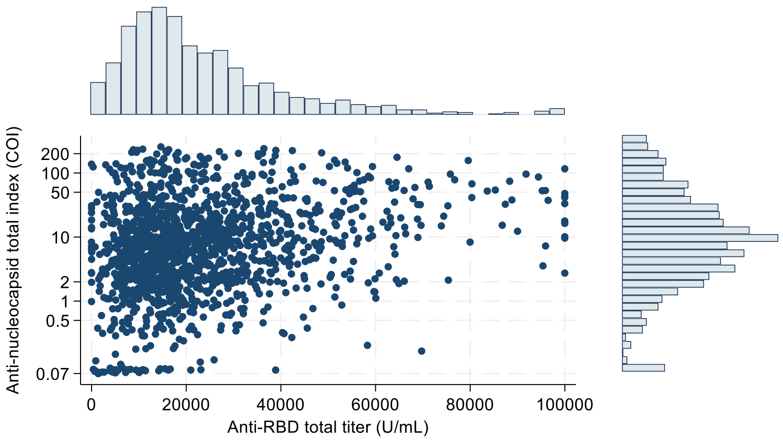

**Abbott**

Spearman's ρ = 0.32 (95%CI: 0.28-0.37)

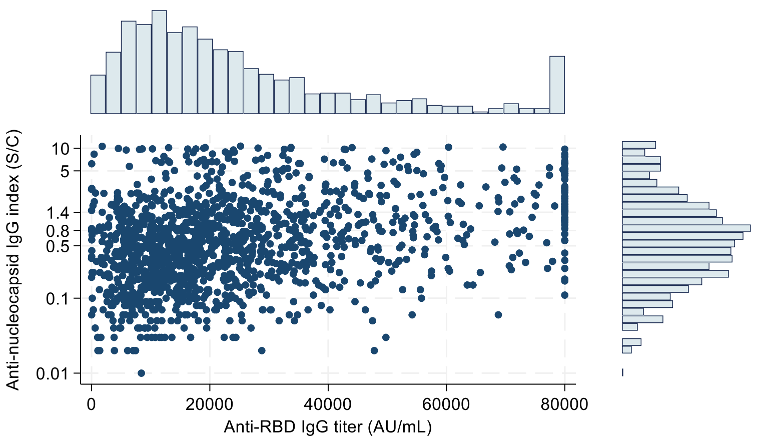

**Sysmex**

Spearman's ρ = 0.34 (95%CI: 0.29-0.38)

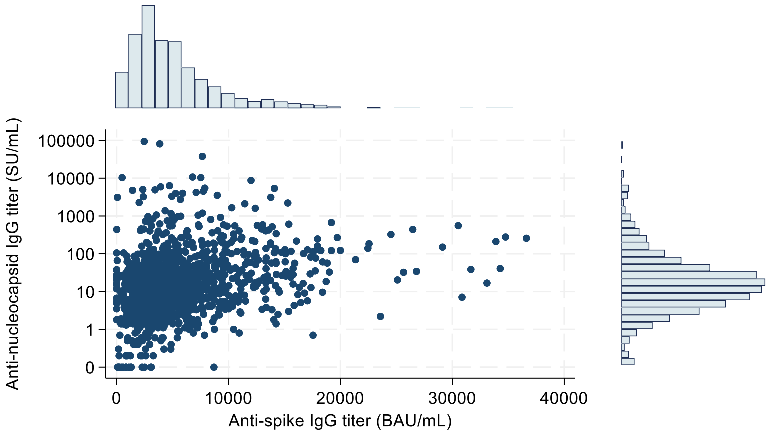

**Supplemental Figure 1.** Scatter plots between anti-nucleocapsid and anti-spike/RBD antibodies with each histogram among previously infected individuals

*Abbreviations*: AU, arbitrary units; COI, cut-off index; IgG, immunoglobulin G; N, nucleocapsid; Q, quartile; RBD, receptor-binding domain; S/CO, signal to cut-off; SU, Sysmex unit.

**Supplemental Table 1.** Association between vaccination status and anti-RBD antibodies by the anti-nucleocapsid antibody status

| Vaccination status | Quartile of anti-RBD total titer (Roche, U/mL) | | | | P for trend |
|---|---|---|---|---|---|
| | S-Q1 | S-Q2 | S-Q3 | S-Q4 | |
| **Infection naïve** | | | | | |
| ≤2-dose | 8.7 | 0.3 | | | <0.01 |
| 3-dose | 23.2 | 10.8 | 6.4 | 1.9 | <0.01 |
| 4-dose | 42.1 | 42.8 | 31.8 | 25.0 | <0.01 |
| 5-dose | 26.0 | 46.1 | 61.8 | 73.1 | <0.01 |
| Interval from last vaccination, days | 294 (189–455) | 200 (177–295) | 182 (172–216) | 175.5 (67–185) | <0.01 |
| **N-Q1: previously infected** | | | | | |
| ≤2-dose | 17.0 | 3.2 | 4.1 | | <0.01 |
| 3-dose | 34.0 | 31.6 | 22.0 | 16.1 | <0.01 |
| 4-dose | 36.2 | 44.2 | 48.0 | 41.9 | 0.58 |
| ≥5-dose | 12.8 | 21.1 | 26.0 | 41.9 | <0.01 |
| Interval from last vaccination, days | 310 (242–523) | 294 (181–510) | 216 (182–304) | 209 (180–298) | <0.01 |
| **N-Q2: previously infected** | | | | | |
| ≤2-dose | 22.2 | 2.4 | 3.0 | 1.0 | <0.01 |
| 3-dose | 36.1 | 36.9 | 25.6 | 15.4 | <0.01 |
| 4-dose | 36.1 | 53.6 | 46.6 | 43.3 | 0.82 |
| ≥5-dose | 5.6 | 7.1 | 24.8 | 40.4 | <0.01 |
| Interval from last vaccination, days | 298 (269–516) | 298 (216–463) | 288 (180–455) | 191.5 (177–295) | <0.01 |
| **N-Q3: previously infected** | | | | | |
| ≤2-dose | 28.6 | 6.1 | 1.8 | 4.3 | <0.01 |
| 3-dose | 23.8 | 43.9 | 32.7 | 23.6 | 0.03 |
| 4-dose | 38.1 | 33.3 | 44.5 | 42.2 | 0.34 |
| ≥5-dose | 9.5 | 16.7 | 20.9 | 29.8 | <0.01 |
| Interval from last vaccination, days | 336 (294–514) | 306 (286–542) | 292.5 (186–462) | 250 (181–309) | <0.01 |
| **N-Q4: previously infected** | | | | | |
| ≤2-dose | 38.5 | 3.4 | 3.1 | | <0.01 |
| 3-dose | 11.5 | 34.5 | 31.6 | 22.9 | 0.69 |
| 4-dose | 34.6 | 48.3 | 45.9 | 44.0 | 0.82 |
| ≥5-dose | 15.4 | 13.8 | 19.4 | 33.1 | <0.01 |
| Interval from last vaccination, days | 285 (181–511) | 296 (194–456) | 298 (220–462) | 284 (181–310) | <0.01 |

Data are presented as percentages for categorical variables and median (interquartile range) for continuous variables.
P for trend was calculated by the Cochran–Armitage test for the categorical variables and by the Jonckheere–Terpstra test for the continuous variables.
Q, quartile; N, nucleocapsid; S, spike

**Supplemental Table 2.** Hazard ratios (95% confidence intervals) for subsequent symptomatic SARS-CoV-2 infection across the baseline anti-nucleocapsid antibody index

| Assays | Infection-naïve | Quartile of N antibodies among previously infected individuals | | | | ***P* for trend** among previously infected individuals |
|---|---|---|---|---|---|---|
| | | **Q1 (lowest)** | **Q2** | **Q3** | **Q4 (highest)** | |
| **Roche (Total N)** | | | | | | |
| COI, median [min–max] | 0.08 [0.07–0.98] | 1.16 [0.07–2.46] | 4.30 [2.48–7.14] | 11.3 [7.15–20.7] | 48.9 [20.8–256] | |
| Cases/Person-days (% [a]) | 169/71901 (23.5) | 33/24958 (13.2) | 7/25897 (2.7) | 11/26564 (4.1) | 5/26677 (1.9) | |
| Model 1 | 1.91 (1.31–2.77) | reference | 0.19 (0.08–0.43) | 0.30 (0.15–0.59) | 0.14 (0.05–0.35) | <0.01 |
| Model 2 | 1.94 (1.32–2.85) | reference | 0.19 (0.08–0.44) | 0.29 (0.15–0.58) | 0.14 (0.05–0.36) | <0.01 |
| Model 3 | 1.57 (1.06–2.32) | reference | 0.19 (0.08–0.44) | 0.33 (0.17–0.66) | 0.17 (0.07–0.43) | <0.01 |
| **Abbott (IgG N)** | | | | | | |
| S/C, median [min–max] | 0.06 [0.01–1.29] | 0.12 [0.01–0.22] | 0.36 [0.23–0.56] | 0.84 [0.57–1.31] | 2.55 [1.32–10.7] | |
| Cases/Person-days (% [a]) | 169/71901 (23.5) | 24/26150 (9.2) | 12/25121 (4.8) | 13/26620 (4.9) | 7/26205 (2.7) | |
| Model 1 | 2.82 (1.84–4.33) | reference | 0.52 (0.26–1.03) | 0.52 (0.26–1.02) | 0.29 (0.12–0.67) | <0.01 |
| Model 2 | 2.92 (1.88–4.53) | reference | 0.53 (0.26–1.06) | 0.52 (0.27–1.03) | 0.29 (0.12–0.68) | <0.01 |
| Model 3 | 2.49 (1.59–3.90) | reference | 0.57 (0.29–1.15) | 0.59 (0.30–1.17) | 0.40 (0.17–0.94) | 0.07 |
| **Sysmex (IgG N)** | | | | | | |
| SU/mL, median [min–max] | 0.1 [0–8.4] | 2.6 [0–5.1] | 8.5 [5.2–13.6] | 21.7 [13.7–34.2] | 97.4 [34.8–94024] | |
| Cases/Person-days (% [a]) | 169/71901 (23.5) | 31/25107 (12.3) | 15/25944 (5.8) | 6/26395 (2.3) | 4/26650 (1.5) | |
| Model 1 | 2.10 (1.43–3.09) | reference | 0.46 (0.25–0.86) | 0.18 (0.08–0.43) | 0.12 (0.04–0.35) | <0.01 |
| Model 2 | 2.15 (1.44–3.19) | reference | 0.45 (0.24–0.84) | 0.18 (0.08–0.44) | 0.12 (0.04–0.35) | <0.01 |
| Model 3 | 1.95 (1.30–2.93) | reference | 0.47 (0.25–0.87) | 0.19 (0.08–0.47) | 0.15 (0.05–0.42) | <0.01 |

Shown are the hazard ratios (95% confidence intervals).
Model 1 was adjusted for age (continuous) and sex (male or female).
Model 2 was additionally adjusted for job (doctors, nurses, allied health professionals, researchers, administrative staff, or others), occupational SARS-CoV-2 exposure risk (low, moderate, or high), body mass index (continuous), comorbid diseases (no or yes), immunosuppression (no or yes), use of tobacco products (no or yes), frequency of alcohol drinking (none, occasional, or weekly/daily drinker), number of households (continuous), number of live-in school-aged children (0, 1, or ≥2), infection prevention score (continuous), spending ≥30 min in the 3Cs without mask (none, 1–5 times, or ≥6 times), and having dinner in a group of ≥5 people for >1 hours (none, 1–5 times, or ≥6 times).
Model 3 was further adjusted for anti-spike/RBD titers measured with the assay of the same company.
[a] Incident rate per 10000 person-day
*Abbreviations*: 3Cs, crowded places, close-contact settings, and confined and enclosed spaces; COI, cut-off index; IgG, immunoglobulin G; N, nucleocapsid; Q, quartile; S/CO, signal to cut-off; SU, Sysmex unit.

**Supplemental Table 3.** Hazard ratios (95% confidence intervals) for subsequent SARS-CoV-2 infection across the baseline anti-nucleocapsid antibody index among participants, excluding non-regular staff (N=2,414)

| **Assays** | **Infection-naïve** | **Quartile of N antibodies among previously infected individuals** | | | | ***P* for trend** among previously infected individuals |
|---|---|---|---|---|---|---|
| | | **Q1 (lowest)** | **Q2** | **Q3** | **Q4 (highest)** | |
| **Roche (Total N)** | | | | | | |
| Cases/Person-days (% [a]) | 168/ 67462 (24.9) | 34/23523 (14.5) | 11/24895 (4.4) | 12/25172 (4.8) | 5/25447 (2.0) | |
| Model 1 | 1.83 (1.27–2.65) | reference | 0.29 (0.14–0.56) | 0.31 (0.16–0.61) | 0.13 (0.05–0.33) | <0.01 |
| Model 2 | 1.85 (1.26–2.71) | reference | 0.28 (0.14–0.56) | 0.31 (0.16–0.60) | 0.13 (0.05–0.34) | <0.01 |
| Model 3 | 1.50 (1.01–2.23) | reference | 0.29 (0.14–0.57) | 0.35 (0.18–0.67) | 0.16 (0.06–0.41) | <0.01 |
| **Abbott (IgG N)** | | | | | | |
| Cases/Person-days (% [a]) | 168/ 67462 (24.9) | 26/24917 (10.4) | 16/23948 (6.7) | 13/24987 (5.2) | 7/25185 (2.8) | |
| Model 1 | 2.61 (1.72–3.95) | reference | 0.64 (0.34–1.19) | 0.49 (0.25–0.95) | 0.26 (0.11–0.61) | <0.01 |
| Model 2 | 2.69 (1.76–4.11) | reference | 0.65 (0.35–1.22) | 0.49 (0.25–0.96) | 0.27 (0.12–0.62) | <0.01 |
| Model 3 | 2.30 (1.49–3.55) | reference | 0.70 (0.38–1.32) | 0.55 (0.28–1.08) | 0.36 (0.15–0.85) | 0.04 |
| **Sysmex (IgG N)** | | | | | | |
| Cases/Person-days (% [a]) | 168/ 67462 (24.9) | 33/24084 (13.7) | 18/24466 (7.4) | 7/25006 (2.8) | 4/25481 (1.6) | |
| Model 1 | 1.99 (1.37–2.89) | reference | 0.53 (0.30–0.94) | 0.20 (0.09–0.45) | 0.11 (0.04–0.32) | <0.01 |
| Model 2 | 2.01 (1.36–2.95) | reference | 0.51 (0.29–0.92) | 0.20 (0.09–0.45) | 0.12 (0.04–0.33) | <0.01 |
| Model 3 | 1.82 (1.22–2.70) | reference | 0.53 (0.30–0.95) | 0.21 (0.09–0.48) | 0.14 (0.05–0.40) | <0.01 |

This analysis done after excluding non-regular staff, including contractors, temporary staff, café staff, shop staff, and part-time registered medical doctors.
Shown are the hazard ratios (95% confidence intervals).
Model 1 was adjusted for age (continuous) and sex (male or female).
Model 2 was additionally adjusted for job (doctors, nurses, allied health professionals, researchers, administrative staff, or others), occupational SARS-CoV-2 exposure risk (low, moderate, or high), body mass index (continuous), comorbid diseases (no or yes), immunosuppression (no or yes), use of tobacco products (no or yes), frequency of alcohol drinking (none, occasional, or weekly/daily drinker), number of households (continuous), number of live-in school-aged children (0, 1, or ≥2), infection prevention score (continuous), spending ≥30 min in the 3Cs without mask (none, 1–5 times, or ≥6 times), and having dinner in a group of ≥5 people for >1 hours (none, 1–5 times, or ≥6 times).
Model 3 was further adjusted for anti-spike/RBD titers measured with the assay of the same company.
[a] Incident rate per 10000 person-day
*Abbreviations*: 3Cs, crowded places, close-contact settings, and confined and enclosed spaces; COI, cut-off index; IgG, immunoglobulin G; N, nucleocapsid; Q, quartile; S/CO, signal to cut-off; SU, Sysmex unit.
